# 18F FDG-PET correlates of motor neuron disease motor variants

**DOI:** 10.64898/2026.02.24.26347019

**Authors:** Boris Deleu, Patrick Dupont, Kato Bracaval, Fouke Ombelet, Frederik Hobin, Nikita Lamaire, Koen Van Laere, Philip Van Damme, Joke De Vocht

**Affiliations:** Department of Neurology, University Hospitals Leuven, Leuven, Belgium; Leuven Brain Institute, KU Leuven, Leuven, Belgium; Laboratory for Cognitive Neurology, Department of Neurosciences, KU Leuven, Leuven, Belgium; Laboratory of Neurobiology, Department of Neuroscience, KU Leuven, Leuven, Belgium; Department of Nuclear Medicine, University Hospitals UZ Leuven, Leuven, Belgium

**Keywords:** Nuclear imaging, 18F FDG PET, motor neuron disease, progressive muscular atrophy, primary lateral sclerosis, amyotrophic lateral sclerosis

## Abstract

While ^18^F-Fluorodeoxyglucose Positron Emission Tomography (FDG-PET) is an established biomarker in amyotrophic lateral sclerosis (ALS), the metabolic correlates of motor neuron disease motor variants remain poorly defined. This is why we investigated patterns of cerebral glucose metabolism across the spectrum of motor neuron disorders (MND), including progressive muscular atrophy (PMA), primary lateral sclerosis (PLS) and amyotrophic lateral sclerosis (ALS).

We retrospectively included 18 PMA, 25 PLS and 43 matched non-hereditary ALS patients according to most recent diagnostic criteria. FDG-PET imaging revealed similar widespread hypometabolism in PMA, as in ALS, whereas PLS showed a more focal motor cortical pattern of hypometabolism.

Despite clinical differences between MND subtypes, PMA and ALS showed similar FDG-PET metabolic patterns, whereas PLS exhibited a more restricted cortical signature in this retrospective study.

## Introduction

Amyotrophic lateral sclerosis (ALS) is a fatal neurodegenerative disorder characterized by involvement of both upper (UMN) and lower (LMN) motor neurons, with variable frontotemporal involvement. It is the most common form of motor neuron disease (MND) (1, 2). Within the MND motor spectrum, primary lateral sclerosis (PLS) is characterized by progressive and isolated UMN degeneration, whereas progressive muscular atrophy (PMA) is defined by isolated LMN involvement (2, 3). Together, PLS and PMA account for up to 10% of MND cases. Clinically, PLS is typically associated with slower disease progression and longer survival compared with ALS and PMA (1, 2). Historically, PLS and PMA were considered restrictive phenotypes (4). However, extra-motor manifestations in PLS and PMA are increasingly recognized (5).

Previous ^18^F-Fluorodeoxyglucose Positron Emission Tomography (FDG-PET) studies to evaluate cerebral metabolism in PLS and PMA are scarce, included small patient cohorts, and their results have been inconsistent (6, 7). Against this background, the aim of our study was to further investigate cerebral metabolic correlates in a larger cohort of PLS and PMA patients.

## Materials and methods

We included 18 patients with PMA, 25 with PLS, 43 matched non-hereditary ALS patients, diagnosed between 2011 and 2021, and 31 healthy controls (8). ALS patients were matched to PMA and PLS cases based on age (±three years), scanner type, and site of onset. The Gold Coast criteria were applied to diagnose ALS patients, as well to evaluate LMN dysfunction in suspected PMA patients (9). PLS patients were included according to most recent consensus criteria (3). The study was approved by the local Ethics and Radiation Safety Committees, and all participants provided written informed consent.

A voxel-based ANCOVA analysis was performed with statistical parametric mapping (SPM12; Welcome Trust Centre for Neuroimaging), implemented in Matlab (R2024b, The MathWorks Inc. Massachusetts, USA). All scans were spatially normalized to Montreal Neurological Institute (MNI) space using a FDG-PET template and non-rigid registration with 16 iterations, in voxels of 2 × 2 × 2 mm. Isotropic Gaussian smoothing with FWHM of 8 mm was performed, followed by proportional scaling applying a relative threshold masking set at 0.8. A voxel-level threshold was set at p_uncorr_<.001, combined with a p_FWE_ <.05 at cluster-level. Details on the PET acquisition and reconstruction can be found in the supplementary materials.

Demographic and clinical characteristics were compared using SPSS 30.0 (SPSS Inc. Chicago, IL, USA), using Kruskal-Wallis H tests for continuous variables and Chi square statistics for categorical variables.

## Results

Demographics and clinical characteristics are reported in Table 1. According to most recent consensus criteria, 18 of the 25 PLS patients were considered definite PLS (no LMN signs >4 years after disease onset). 7 patients were considered as probable PLS (no LMN signs 2-4 years after disease onset) (3). MND phenotypes differed significantly in diagnostic delay ((χ2(2) = 27.0635), p <.001), survival (χ2(2) = 69.033, p <.001) and sex (χ2(2) =9,198, p =.010) (Table 1). The Kaplan-Meier survival curves are presented in Figure 1.

**Table 1:** Demographic and clinical characteristics. ALS, Amyotrophic lateral sclerosis; ALSFRS-R, Revised ALS Functional Rating Scale; FDG-PET: ^18^F-Fluorodeoxyglucose Positron Emission Tomography; IQR, Interquartile range; M, months; n, number; PLS, Primary Lateral Sclerosis; PMA, Progressive Muscular Atrophy; SD, Standard deviation; Y, years; ΔFS, ALSFRS-R progression slope. *: data available in 100% of ALS, 84% of PLS and 78% of PMA cases.

|  | <u>ALS</u><br>(n= 43) | <u>PLS</u><br>(n= 25) | <u>PMA</u><br>(n= 18) | <u>Controls</u><br>(n= 31) |
| --- | --- | --- | --- | --- |
| Sex (Male)<br>(p-value = 0.01) | 28 (65%) | 11 (44%) | 16 (89%) | 16 (52%) |
| PET/CT |  |  |  |  |
| • Hirez | 16 (37%) | 8 (32%) | 5 (28%) | - |
| • Truepoint | 16 (37%) | 12 (48%) | 7 (39%) | 31 (100%) |
| ECAT HR+ | 11 (26%) | 5 (20%) | 6 (33%) | - |
| Site of Onset (Spinal) | 33 (77%) | 16 (64%) | 17 (94%) | - |
| Deceased | 41 (95%) | 3 (12%) | 13 (72%) | - |
| **<u>Mean ± SD [Range]</u>** |  |  |  |  |
| Age at symptom onset (Y) | 61 ± 9<br>[42 - 83] | 58 ± 10<br>[42 - 78] | 60 ± 11<br>[38 - 77] |  |
| Age at FDG-PET imaging (Y) | 62 ± 9<br>[43 - 84] | 62 ± 9<br>[46 - 83] | 62 ± 10<br>[44 - 79] |  |
| Diagnostic delay (M)<br>(p-value < .001) | 13 ± 8<br>[3 - 36] | 38 ± 28<br>[7 - 108] | 25 ± 19<br>[12 - 87] |  |
| Clinical follow up from symptom onset (M) | 42 ± 22<br>[12 - 86] | 101 ± 67<br>[24 - 252] | 61 ± 43<br>[22 - 156] |  |
| Survival (M) + IQR<br>(p-value < .001) | 42 ± 22<br>[12 - 86]<br>[Q1=23;<br>Q3=60] | 132 ± 76<br>[38 - 360]<br>[Q1 and Q3<br>not reached] | 98 ± 52<br>[25 - 188]<br>[Q1=58;<br>Q3not<br>reached] |  |
| ALSFRS-R (closed to FDG-PET) * | 40 ± 4<br>[24 - 47] | 38 ± 8<br>[19 - 46] | 40 ± 6<br>[29 - 48] |  |
| ΔFS* | 0.77 ± 0.56<br>[0.08 - 2.00] | 0.26 ± 0.19<br>[0.04 - 0.60] | 0.36 ± 0.36<br>[0 - 1.08] |  |

**Figure 1.**
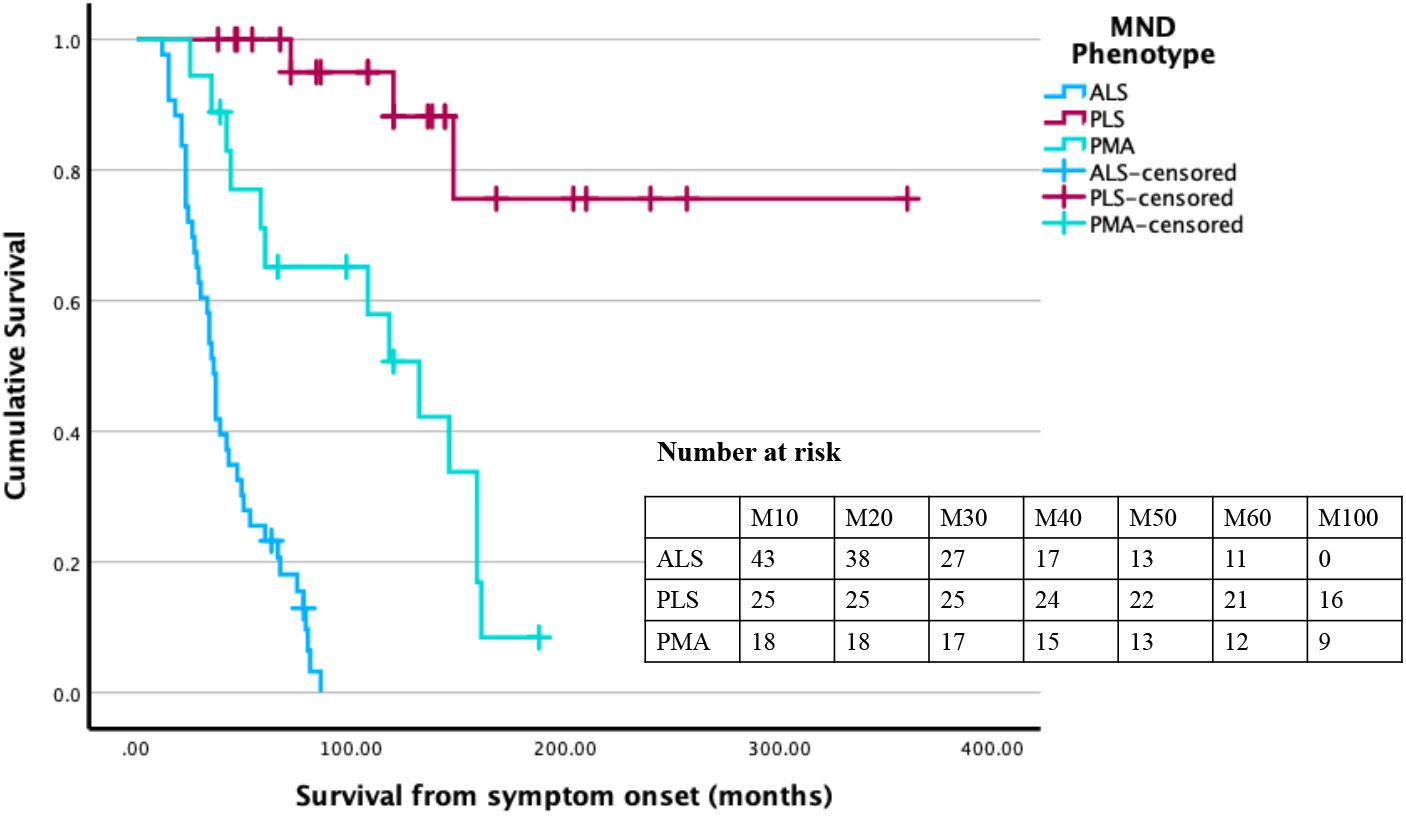
Kaplan-Meier survival curve of the different MND phenotypes. ALS, Amyotrophic lateral sclerosis; M, month; MND, motor neuron disease; PLS, Primary Lateral Sclerosis; PMA, Progressive Muscular Atrophy.

Direct comparison between MND phenotypes, revealed clusters of relative hypometabolism in (pre)frontal and occipito-temporal regions in ALS and (pre)frontal regions in PMA when compared to PLS (Figure 2). However, no significant differences were found when comparing PMA and ALS patient groups.

**Figure 2.**
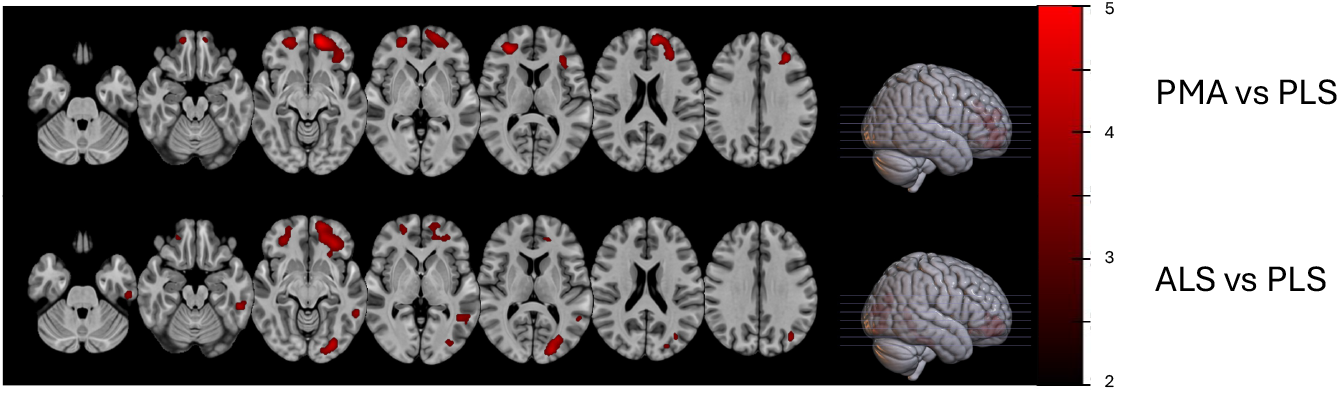
Voxel-based comparison of the different MND phenotypes. Surface projections of areas with relative hypometabolism (red) across motor neuron disease phenotypes, while correcting for age, sex and scanner, P_uncorr_ <.001, combined with a P_FWE_ <.05 at cluster level. ALS, Amyotrophic lateral sclerosis; PLS, Primary Lateral Sclerosis; PMA, Progressive Muscular Atrophy.

A follow-up analysis between MND phenotypes and a cohort of healthy controls revealed widespread hypometabolism in the (pre)frontal, parietal and temporal regions, together with relative hypermetabolism in cerebellar areas in ALS. PMA patients exhibited a similar pattern of relative hypometabolism to that observed in ALS, without areas of hypermetabolism. In PLS, comparison with controls revealed focal hypometabolism around the motor cortex, as well as relative hypermetabolism in occipital, parieto-temporal, and frontal regions (figure 3).

**Figure 3.**
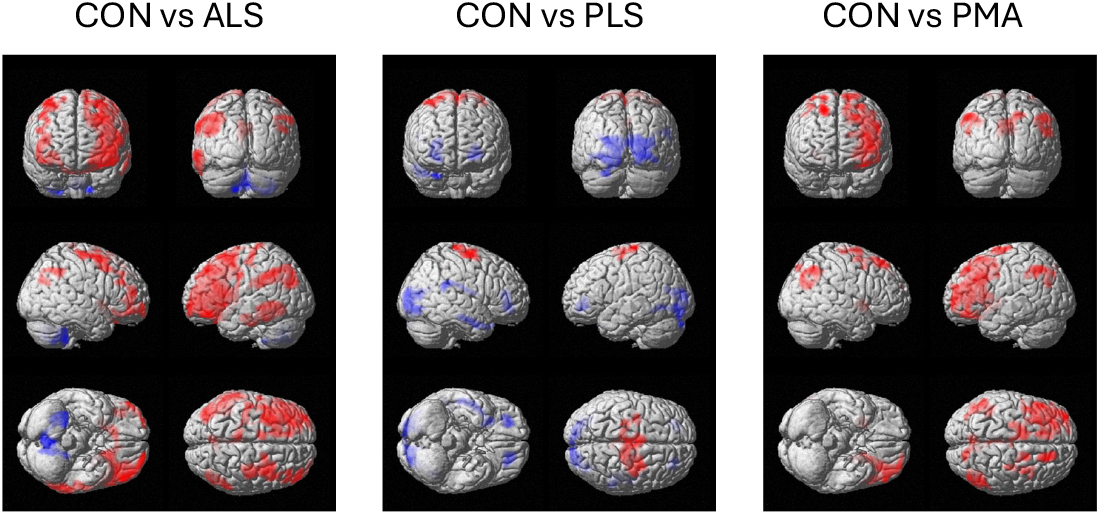
Voxel-based comparison of the different MND phenotypes with healthy controls. Surface projections of areas with relative hypometabolism (red) and relative hypermetabolism (blue), while correcting for age and sex, P_uncorr_ <.001, combined with a P_FWE_ <.05 at cluster level. As healthy controls were collected on Truepoint PET/CT, we only considered patients that were also scanned on Hirez or Truepoint PET/CT. ALS, Amyotrophic lateral sclerosis; PLS, Primary Lateral Sclerosis; PMA, Progressive Muscular Atrophy

## Discussion

Despite distinct clinical differences regarding UMN involvement, FDG-PET revealed a marked similarity in cerebral hypometabolism between PMA and ALS, in comparison to healthy controls. This finding has previously been reported in patients with PMA, although its interpretation was limited by small sample size at the time (6). This metabolic similarity could indicate shared upstream pathophysiological mechanisms, despite phenotypic differences during clinical examination, indicating that PMA and ALS are phenotypes of the same MND spectrum. In contrast, PLS exhibited a more confined metabolic pattern, with less severe extra-motor involvement, when compared to PMA and ALS cases.

Our clinical observations corroborate earlier findings, as patients with PLS in our study showed a significantly longer survival compared with patients with ALS and PMA, as well as a markedly longer diagnostic delay, reflecting the typically slower disease course and diagnostic challenges associated with this phenotype (1, 2). In addition, PMA predominantly affected male patients, consistent with earlier epidemiological reports (10).

This retrospective study had several limitations. The most important one for interpretation of the results is the fact that it is unclear what the distribution of cognitive and behavioural changes was in our ALS, PLS and PMA cohorts, and its relation to FDG uptake, as we did not subject most patients to a cognitive screening. Emerging evidence suggests that such cognitive and behavioral changes may also be observed in PMA and PLS, thereby challenging the concept of restricted phenotypes (5).

## Data Availability

All data produced in the present study are available upon reasonable request to the authors.

## Acknowledgements

PVD is supported by grants from KU Leuven (C1 - C14/22/132), Opening the Future Fund (KU Leuven), the Fund for Scientific Research Flanders (FWO Vlaanderen n° G026125N, G073222N), Target ALS (FS-2024-ESC-S1), the ALS Liga België and the KU Leuven funds “Een Hart voor ALS” and “Laeversfonds voor ALS Onderzoek”. PVD holds a fundamental clinical investigatorship of KU Leuven.

J.D.V. is supported by FWO (Postdoctoral Fellowship, 12AQF24N).

## Notes

### Competing Interest Statement

The authors have declared no competing interest.

### Funding Statement

This study did not receive any funding.

### Author Declarations

The study was approved by the Ethics and Radiation Safety Committees at UZ/KULeuven.

