## Supplementary materials for "18F FDG-PET correlates of motor neuron disease motor variants"

*Image analysis: PET Acquisition and Reconstruction*

All subjects underwent a diagnostic FDG-PET scan. All subjects fasted at least 6 hours before an [$^{18}$F]-FDG bolus was injected intravenously under standard conditions: lying in a dimly lit, quiet room with eyes and ears open. FDG-PET scans were acquired using an ECAT HR+ PET camera (Siemens Healthcare, Erlangen, Germany) operated in 3D mode, a Biograph 16 HiRez PET-CT camera (Siemens Healthcare, Erlangen, Germany) or Siemens Biograph 40 'TruePoint' PET/CT. Thirty minutes after injection, images were acquired dynamically for 30 minutes on the ECAT HR+ (6 frames, 5 minutes per frame) and for 15-20 minutes on the HiRez and Truepoint (listmode). During the acquisition, the patients' head was immobilized with a vacuum pillow.

On the ECAT HR+, images were corrected for attenuation using a $^{68}$Ge rod source and were reconstructed using 3-dimensional filtered back projection with a Hanning post-filter. Before the PET scan, a low-dose CT scan was acquired to generate a CT-based attenuation map, on the HiRez or Truepoint. PET images on either PET/CT were reconstructed using ordered-subset expectation maximization (OSEM), followed by post-reconstruction smoothing with a 2-3 mm Full Width Half Maximum (FWHM) Gaussian kernel. Scanner-specific PET image reconstruction procedures have been described in detail by Tang et. al. (2025).

### Supplementary tables

| | Cluster level | | Peak level | | MNI coordinates (mm) | | | Region |
|---|---|---|---|---|---|---|---|---|
| | $p_{FWE}$ | Cluster size | T | Z | x | y | z | |
| **ALS < PLS** | .001 | 848 | 4.32 | 4.08 | -20 | -90 | 12 | Occipital Pole |
| | | | 4.27 | 4.04 | -30 | -82 | -10 | Lateral Occipital Cortex, superior division |
| | | | 4.05 | 3.85 | -28 | -76 | 10 | Lateral Occipital Cortex, superior division |
| | <.001 | 1120 | 4.26 | 4.03 | -30 | 42 | -12 | Frontal Pole |
| | | | 4.22 | 4.00 | -16 | 58 | -8 | Frontal Pole |
| | | | 3.60 | 3.46 | -26 | 24 | -12 | Frontal Orbital Cortex |
| | .014 | 503 | 4.24 | 4.01 | -52 | -36 | -22 | Middle Temporal gyrus, posterior division |
| | | | 3.92 | 3.74 | -56 | -24 | -26 | Inferior Temporal gyrus, posterior division |
| | | | 3.78 | 3.62 | -58 | -46 | -10 | Inferior Temporal gyrus, temporooccipital part |
| | 0.043 | 367 | 3.95 | 3.76 | 28 | 44 | -12 | Frontal Pole |
| | | | 3.59 | 3.45 | 20 | 54 | 0 | Frontal Pole |
| **PMA < PLS** | <.001 | 1034 | 4.99 | 4.64 | -14 | 54 | -10 | Paracingulate gyrus |
| | | | 3.67 | 3.52 | -34 | 36 | -8 | Frontal Orbital Cortex |
| | 0.002 | 782 | 4.53 | 4.26 | 30 | 46 | 10 | Frontal Pole |
| | | | 4.24 | 4.01 | 16 | 46 | -16 | Frontal Pole |
| | | | 4.14 | 3.93 | 16 | 56 | -18 | Frontal Pole |
| | 0.003 | 717 | 4.52 | 4.25 | -10 | 54 | 20 | Frontal Pole |
| | | | 4.15 | 3.94 | -32 | 34 | 30 | Middle Frontal gyrus |
| | | | 3.97 | 3.78 | -28 | 40 | 22 | Frontal Pole |

STable 1: Significant clusters of relative hypometabolism and hypermetabolism when comparing different MND phenotypes. ALS, Amyotrophic lateral sclerosis; FWE, Family-wise-error; MNI, Montreal neurological Institute; PLS, Primary Lateral Sclerosis; PMA, Progressive Muscular Atrophy.
